# Multimodal cell-free DNA whole-genome analysis combined with TET-Assisted Pyridine Borane Sequencing is sensitive and reveals specific cancer signals

**DOI:** 10.1101/2023.09.29.23296336

**Authors:** Dimitris Vavoulis, Anthony Cutts, Nishita Thota, Jordan Brown, Robert Sugar, Antonio Rueda, Arman Ardalan, Flavia Matos Santo, Thippesh Sannasiddappa, Bronwen Miller, Stephen Ash, Yibin Liu, Chun-Xiao Song, Brian Nicholson, Helene Dreau, Carolyn Tregidgo, Anna Schuh

## Abstract

The analysis of circulating tumour DNA (ctDNA) promises to extend current tissue-specific cancer screening programmes to multi-cancer early detection and measurable disease monitoring to solid tumours using minimally invasive blood draws (liquid biopsies). Most studies so far have focussed on using targeted deep sequencing to detect the low-abundance, fragmented ctDNA. A few studies have integrated information from multiple modalities using shallow 1× WGS. Here, we developed an integrated bioinformatics pipeline for ctDNA detection based on whole genome TET-Assisted Pyridine Borane Sequencing (TAPS) of plasma samples sequenced at 80× or higher. We conducted a diagnostic accuracy study in a case-control cohort of patients presenting to the UK National Health Service’s (NHS) primary care pathway with non-specific symptoms of cancer, who either did not have cancer or who were subsequently diagnosed with cancer and referred to surgery with curative intent. TAPS is a base-level-resolution sequencing methodology for the detection of 5-methylcytosines and 5-hydro-methylcytosines. Unlike bisulfite-sequencing, the current established method for mapping epigenetic DNA modifications, TAPS is a non-destructive methodology, which only converts methylated cytosines and preserves DNA fragments over 10 kilobases long, thus opening the possibility of simultaneous methylome and genome analysis on the same sequencing data. The proposed methodology combines copy number aberrations and single nucleotide variants with methylation calls from TAPS-treated plasma from patients with Stage 1-4 colorectal (n=36), oesophageal (n=8), pancreatic (n=6), renal (n=5), ovarian (n=4) and breast (n=2) cancers. Plasma samples from 21 confirmed non-cancer controls were used for data denoising, while plasma samples from 9 additional age-matched healthy controls were further used to establish the minimum level of detection. Copy number aberrations, single nucleotide variants, and methylation signals were independently analysed and combined in sample-specific scores, which quantify the levels of plasma ctDNA. Matched tumour samples were used for validation, not for guiding the analysis, imitating an early detection scenario. The detection threshold was set such that specificity was 100%, resulting in sensitivity of 85.2%. In silico experiments on high-fidelity synthetic data suggest excellent discriminatory capacity (AUC > 80%) at ctDNA fractions as low as 0.7%. Furthermore, we demonstrate successful tracking of tumour burden post-treatment and ctDNA shedding in precancerous adenomas in patients with colorectal cancer in the absence of a matched tumour biopsy. In summary, we developed and validated a pipeline for interrogating liquid biopsies using TAPS 80× or higher WGS that is ready for in-depth clinical evaluation both in multi-cancer screening of high-risk individuals and multi-cancer measurable disease monitoring.

## Introduction

Earlier cancer detection has the potential to improve patient outcomes^1^. Current screening programmes around the world are limited to specific cancers (cervical, breast, colorectal, lung, prostate) that together make up less than 30% of all cancer diagnoses. Importantly, uptake of screening, especially of invasive procedures, depends on acceptance in the population. Multi-cancer early detection (MCED) using minimally invasive liquid biopsies therefore holds great promise. However, it also poses challenges from the inherent false positive rate in asymptomatic individuals caused by the low prevalence of cancer even in enriched risk groups^2,3^. Demonstration of efficacy, safety and cost-effectiveness therefore requires large, randomised studies^4^.

Targeting early cancer detection to high-risk individuals presenting to primary care with non-specific symptoms of cancer represents an alternative approach^5,6^. Symptoms have poor predictive value for cancer in low prevalence settings such as primary care, where the tools for risk stratification remain limited^7,8^. For example, in the UK National Health Service (NHS), only 7.0% of 2.07 million referrals from general practitioners (GPs) to secondary care in 2020/2021 resulted in a cancer diagnosis, accounting for 55% of cancer diagnoses that year^9^. Specifically for patients with non-specific symptoms, 241 cancers were diagnosed following 2961 referrals, with a conversion rate of 8.1% spread across multiple cancer sites^10^.

Specialised rapid diagnostic clinics to investigate for cancer at multiple sites and to explain symptoms^10,11^ have already been introduced in some countries. Now, better tools that assist GPs with identifying patients with a high clinical suspicion of cancer are urgently needed^12–15^.

Liquid biopsies as one such potential initial triage tool have recently been investigated in the UK National Health Service (NHS)^6^. The Galleri GRAIL technology that exploits cancer-specific methylation signals, resulted in an overall sensitivity of 66.3% (61.2-71.1%), and specificity of 98.4% (98.1-98.8%). Seventy-nine out of 5461 individuals had a cancer signal without subsequent cancer diagnosis. Sensitivity increased with increasing age and cancer stage, from 24.2% (16.0-34.1%) in Stage I to 95.3% (88.5-98.7%) in Stage IV. Where a cancer signal was detected among cancer patients the MCED test’s prediction of the site of origin was accurate in 85.2% (79.8-89%) of cases.

Other technologies to interrogate liquid biopsies are currently under investigation^16^ in case-control studies to further improve sensitivity while keeping specificity as close to 100% as possible. Most deploy targeted deep sequencing of circulating tumour DNA (ctDNA) and limit detection to the most common types of cancers and cancer-specific, acquired SNVs; some combine this information with targeted epigenetic analysis and protein markers, to improve sensitivity across a wider range of different tumours^2,17,18^.

Few studies so far propose interrogating liquid biopsies by shallow whole genome sequencing (WGS) for either cancer-specific, acquired CNAs^19^ or for specific ctDNA fragment attributes^20,21^ as an alternative to targeted deep sequencing of SNVs, thus supporting the notion that breadth can replace depth^22^. So far, very little data on few patients undergoing treatment has been published on integrating information from multiple modalities using deeper 30×-100× WGS^22–27^.

Tissue-type specific methylation patterns are used to detect cancer signals and tissue of origin (TOO) in ctDNA and are historically derived from bisulfite sequencing following a similar approach to that employed in the Galleri assay. However, bisulfite treatment destroys up to 80% of the already scant ctDNA and hence reduces sensitivity significantly^28^. Besides, it converts the 95% of unmethylated cytosines to thymines, thereby destroying the genetic code for alignment and making SNV calling impossible. Alternative methodologies based on enzymatic conversion have been explored^29–31^.

TET-Assisted Pyridine Borane Sequencing (TAPS) is a base-level-resolution sequencing methodology for the detection of 5-methylcytosines and 5-hydro-methylcytosines^29,32^. Unlike bisulfite-sequencing, the currently established method for mapping epigenetic DNA modifications, TAPS is a non-destructive methodology, which employs a combination of TET enzyme with borane to exclusively convert the 5% of methylated cytosines, thus preserving the genetic code and opening the possibility of simultaneous methylome and genome analysis on the same sequencing data.

Finally, for large scale implementation, other test attributes in addition to sensitivity and specificity, such as quantity of plasma required, turn-around time (TAT) and the scalability of the laboratory processes (in particular, ease of automation and robustness of library preparation for clinical accreditation) are critical. In the NHS Genomic Medicine Service, and in many cancer centres across the world, WGS is already routinely performed for genetic analysis of tumour samples. Expanding the WGS repertoire to indications that require ctDNA analysis would therefore be feasible.

Here, we therefore opted to distinguish true cancer signals in ctDNA from non-cancer noise using integrative multi-modal TAPS WGS in a case control series of UK NHS patients presenting to primary care who either had non-specific symptoms of cancer but did not have cancer or who were subsequently diagnosed with cancer and referred to surgery with curative intent and we compared results from these cohorts with those of healthy age-matched controls.

## Methods

### Study design and clinical cohorts

We conducted a diagnostic accuracy study using a case-control design (Fig. 1; Table 1). Patients were eligible for recruitment if they were aged 18 years or above, willing and able to give informed consent for participation in the study and were referred for urgent investigation for a possible gynaecological, lower GI or upper GI or renal cancer or to a rapid diagnostic centre (RDC) with non-specific symptoms that might be due to cancer. Referral criteria for each pathway were as summarised in the National Institute for Health and Care Excellence (NICE) Guideline 12 Suspected cancer: recognition and referral (NG12) (Supplementary Material)^33^. Patients could not enter the study if they had a history of invasive or haematological malignancy diagnosed within the preceding 3 years or if they were taking cytotoxic or demethylating agents that might interfere with test performance. Only samples of patients with confirmed cancer diagnosis who were advised to undergo surgery with curative intent were selected. For non-cancer controls, only samples of patients referred with non-specific symptoms of cancer who had not developed cancer within the subsequent two years were chosen for signal-to-noise control. Asymptomatic age and sex matched subjects served as negative controls.

**Figure 1:**
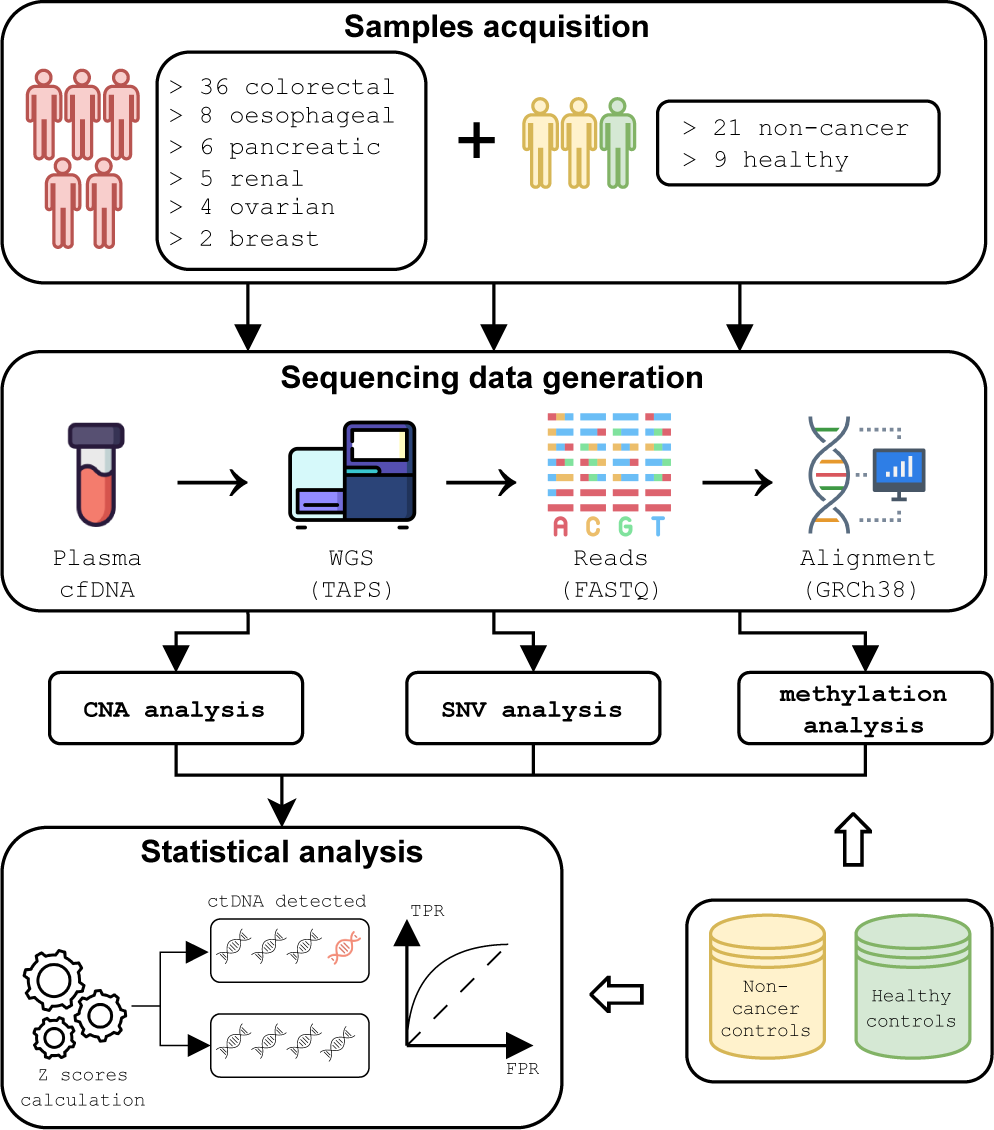
Overview of the study. We conducted a diagnostic accuracy study using a case-control design. We recruited a cohort of UK NHS patients presenting to primary care, who either had non-specific symptoms of cancer but did not have cancer, or who were subsequently diagnosed with cancer and referred to surgery with curative intent. After collection of plasma samples, we conducted whole-genome sequencing at 80× or higher using TET-Assisted Pyridine Borane Sequencing (TAPS), aligned the generated reads against the human genome (GRCh38), and conducted analyses of copy number aberrations, methylation modifications, and somatic point mutations and indels, which included efficient denoising using non-cancer controls. By integrating the analyses from all three data modalities, we generated sample-specific scores for the quantification of plasma ctDNA burden, which was used for cancer detection and post-treatment disease tracking. Matched tumour biopsies, if available, were used for validation, not for guiding the analysis.

**Table 1:**
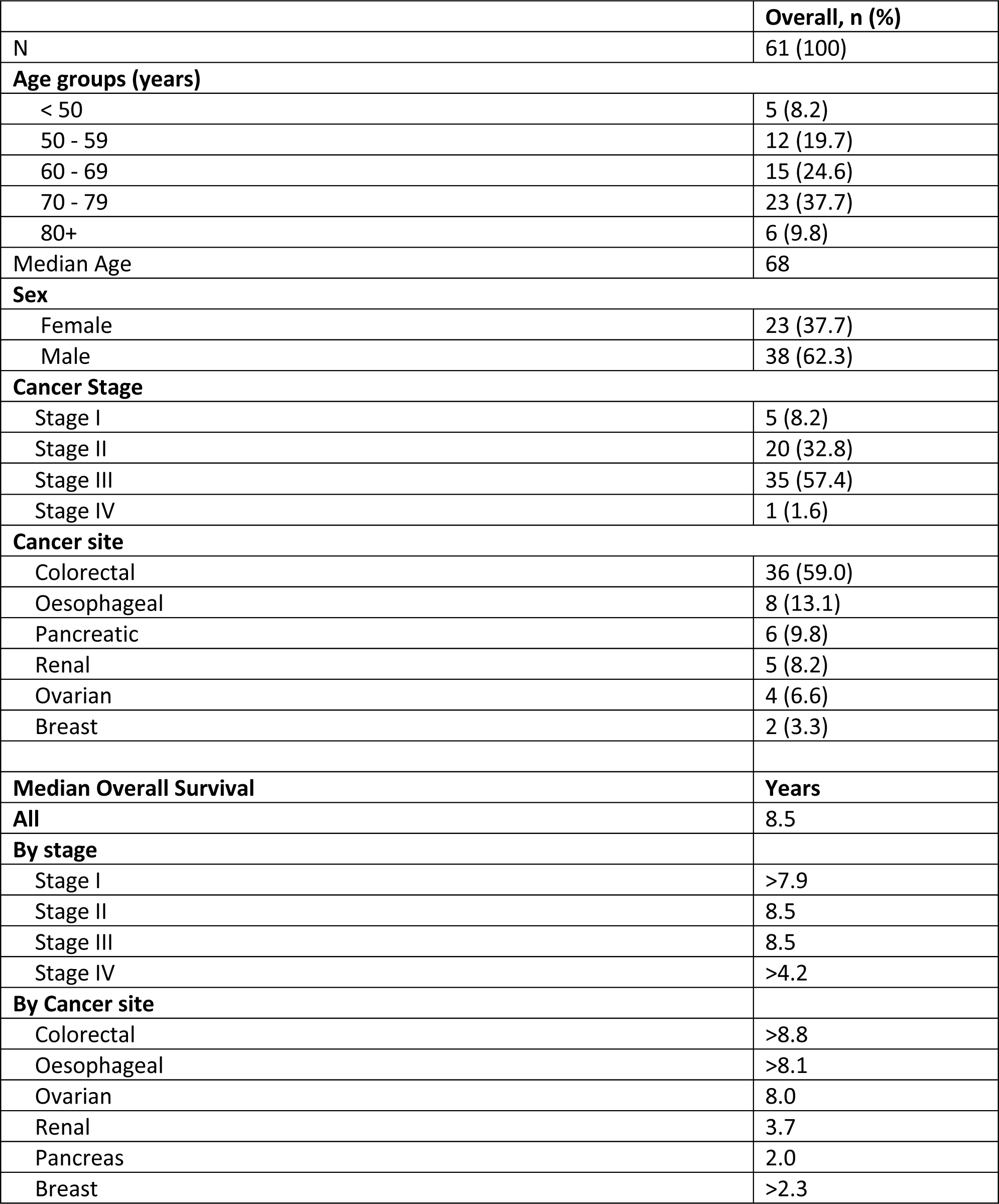
Clinical characteristics of the 61 patients used in this study. Subjects from this cohort presented to the NHS primary care pathway with non-specific symptoms of cancer, who either did not have cancer or who were subsequently diagnosed with cancer and referred to surgery with curative intent.

For measurable disease monitoring, we chose a cohort of 10 patients with colorectal cancer undergoing surgery with or without adjuvant chemotherapy. Post-surgery samples were taken from all patients at least 6 weeks after surgery. For 5 patients, additional follow-up samples were obtained. In total, WGS data from ctDNA was available from 26 samples.

### Ethics

The National Health Service (NHS) Health Research Authority South Central—Oxford C Research Ethics Committee approved this study, and all research was performed in accordance with relevant regulations and guidelines and with the Declaration of Helsinki. Written informed consent was obtained for patients recruited into the 100,000 Genomics England (GEL) pilot study and the rapid diagnostic clinic research pathway in Oxford called Suspected CANcer (SCAN) according to Oxford Radcliffe Biobank (ORB) guidelines (Oxford C Research Ethics Committee Number: 19/SC/0173). For the healthy controls, blood was received from Cambridge Bioscience Human Blood Products Supply Service, where comprehensive informed written consent was provided in accordance with UK ethics and consent regulations.

### Clinical sample collection

Blood samples were collected into K_2_EDTA tubes in all cases for germline DNA (gDNA) extraction from normal peripheral blood leucocytes. For GEL pilot patients, blood was collected into additional K_2_EDTA tubes and processed within 4 hours for plasma extraction. For all other cases, blood was collected into PAXgene Blood ccfDNA tubes and processed within 72 hours of venepuncture. Blood samples were centrifuged for 10 min at 1600 g to separate the plasma, which was subsequently aliquoted into fresh tubes and underwent a second centrifugation for 10 min at 4500 g. The plasma supernatants were stored in aliquots at -80°C prior to extraction. Tissue samples were obtained during surgical resection and Fresh Frozen (FF) until required for extraction.

### DNA Extraction

cfDNA was extracted using the QIAamp circulating nucleic acid kit (Qiagen), according to the manufacturer’s instructions. Input plasma volumes ranged from 4 ml to 25 ml per sample, where the maximum input volume per extraction was 5 ml. Where more than one extraction was carried out per patient, extracted cfDNA was pooled. DNA was first eluted into 30 µl of buffer AVE and subsequently a second elution of 30 µl to maximise yield. gDNA was extracted using the QIAamp Blood Mini kit (Qiagen), according to the manufacturer’s instructions. FF tissue samples initially underwent disruption and homogenization using a TissueRupter and DNA tumour DNA (tDNA) was extracted using the QIAamp Allprep Mini kit (Qiagen), according to the manufacturer’s instructions. All samples were quantified using the Qubit Fluorimeter (ThermoFisher) high sensitivity dsDNA assay.

### Library Preparation and Sequencing

A methylated control (control A) and an unmethylated control (control B) were spiked-in to the extracted DNA. 1 µg of GL and FF DNA spiked with controls were fragmented to 450 bp **Error! Reference source not found.**using the M220 focused-ultrasonicator (Covaris). GL and FF DNA were size-selected for 300-500 bp fragments using 0.55X followed by 0.75X ratios of sample to Agencourt AMPure XP Beads (Beckman Coulter). 50 ng of cfDNA were spiked with the same two controls previously fragmented to 150bp. End repair and adapter ligation adjusted with a 1:10 dilution of adapters for cfDNA, were performed using the KAPA HyperPrep Kit (KAPA Biosystems), according to the manufacturer’s recommendations. Post adapter ligation, an AMPure bead purification step was done using 0.8X ratio of beads to sample and DNA eluted in buffer EB. DNA oxidation used reagents and protocols supplied by Exact Sciences Innovation. Oxidation was followed by an incubation with Proteinase K (0.8U) (New England Biolabs (NEB)) at 50°C for 30 minutes, stopping the oxidation. This was followed by an AMPure bead clean-up using a 1.8X bead to sample ratio. Reduction was completed using borane compound and other reagents and protocols supplied by Exact sciences Innovation. Subsequently, libraries were purified using AMPure XP beads at a 1.8X bead to sample ratio. A first extension was completed with an enzyme mix from Exact Sciences Innovation before library amplification was performed using KAPA HiFi HotStart Uracil+ ReadyMix Kit (KAPA Biosystems), according to the manufacturer’s instructions, with the exception that i5 and i7 NEBNext Multiplex Dual Oligos for Illumina (NEB) were used. GDNA and tDNA libraries underwent 4 PCR cycles, while 6 PCR cycles were performed on cfDNA libraries. Final libraries were purified using AMPure beads at a 1X ratio and DNA was eluted into buffer EB. To assess the conversion of 5mC, amplification of a 194bp DNA fragment from the methylated lambda control was performed, using the primers F_5’-GCTGGGGAACTACAGGCT-3’ and R_5’-AGAACCAGAACTCAAACTGTAC-3’ (Integrated DNA Technologies). A PCR mastermix was made containing 1 µl 10× Standard Taq Buffer, 0.5 µl 10mM dNTPs, 0.5 µl 10mM primers, 0.25 µl Taq polymerase, 2 ng DNA template and the final volume made up to 50 µl with nuclease-free water. Thirty-five cycles of PCR were carried out (1 cycle of 95 °C 30 s, 35 cycles of 95 °C, 48°C 30 s, 68°C 1 s, followed by 1 cycle of 68°C 5 s). Once complete, a restriction-enzyme digest mix was set up containing 1 µl 10X Cutsmart buffer, 0.2 µl TaqI, 5 µl PCR product and made up to 10 µl with nuclease water. This mix was incubated at 65°C for 30 minutes and the products analysed using agarose gel electrophoresis. Shallow sequencing runs for initial quality metrics were performed using a MiniSeq (Illumina). Paired-end (2×150bp) runs utilised the MiniSeq High Output Reagent Kit (300-cycles) aiming for a depth of at least 0.4x. Whole genome sequencing was performed on a NovaSeq 6000 (Illumina) (paired-end reads–2×150) aiming at 30× sequencing depth for GL and at least 80× for FF and cfDNA on an S4 flowcell (Illumina).

### Coverage signal extraction and denoising

Each cfDNA BAM file was segmented into 1000bp-long nonoverlapping bins, and the number of alignments in each bin was counted and reported using `bedtools intersect v2.30.0`. Only properly paired non-duplicated reads with high mapping quality (MAPQ > 30) were considered. These genome-wide counts represent the raw coverage signal, which was then brought into a state appropriate for statistical analysis through several pre-processing steps, as follows. First, we subtracted from the raw signal all ENCODE blacklisted regions v2 and all difficult regions from the Genome-In-A-Bottle project. In a subsequent filtering step, we removed bins with mappability score less than 50%, as well as bins with excessively high or low GC content, as these indicate the presence of potential artefacts. In the following step, we normalised the coverage signal in each bin by dividing by the genome-wide median coverage, and then taking the log_2_ value of the resulting ratio. Each bin was then annotated with its median GC content and mappability score. Since the coverage signal in each bin is dependent on its GC content and mappability, it must be corrected for any bias introduced by these two variables. For this purpose, we used a Generalised Additive Model (GAM) to describe the coverage signal as a function of GC content and mappability. The choice of a GAM allows us to model the coverage without assuming any particular form for the functions that model its dependence on GC content and mappability. In `R v4.1.3`, this is done using the function `gam` from package `mgcv v1.8-40`, as follows: `gam(y∼s(GC, bs=’cs’) + s(MAP, bs=’cs’))`. In this code snippet, *y* is the vector of normalised coverage values across all bins, while GC and MAP are the corresponding vectors of GC content and mappability scores. The coverage *y* is corrected by subtracting its dependencies on GC and MAP, as estimated using the above procedure. Finally, the de-biased coverage signal in each bin is normalised again by subtracting the genome-wide average. The filtered and de-biased coverage for all plasma cfDNA samples obtained from the previous steps is further denoised, using the identically pre-processed non-cancer SCAN cfDNA samples for characterising the systematic background noise. These SCAN samples encapsulate unwanted variance, which we wish to remove from both healthy controls and cancer samples. This is achieved in two stages. We start by collecting the non-cancer SCAN samples in a matrix *M* with *m* columns (equal to the number of SCAN samples) and *n* rows (equal to the total number of bins), where *m* is typically much smaller than *n*. We factorise this matrix using its singular value decomposition, *M* = *U*Σ*V*^*T*^, where Σ is an *m* × *m* square matrix with the singular values along its main diagonal in decreasing order, *U* is an orthogonal *n* × *m* matrix with the corresponding left singular vectors as its columns, and *V* is also an orthogonal *m* × *m* matrix with the corresponding right singular vectors as its columns. If the coverage signal for a particular sample is *y*, the systematic background noise is given by the product *UU*^*T*^*y*, which is subsequently subtracted from *y*. In the second stage, the coverage signal is further denoised through application of the Savitzky-Golay filter^34^, a digital filter that smooths the data without distorting the underlying signal. In `R v4.1.3`, this can be achieved using the function `sgolayfilt` from the R package `signal v0.7-7`. We use a filter of order 3 with a filter length of 1000 bins (10^6^ base pairs). After smoothing, the denoised signal is again normalised by subtracting the genome-wide median.

### Somatic variant callin

An in-house (ExactSciences) TAPS methylation caller, `TAMER v0.34.0`, was used to call methylation from the cfDNA and the germline BAM files. The TAMER algorithm recognizes TAPS changes (mC>T) and differentiates these from C>T variants or variants resulting in a methylated C>T. It can also indicate the co-location of methylation change and a variant. Upon TAMER analysis, a cfDNA and a germline methylation VCF was generated. For both the cfDNA and germline, the TAPS methylation calls were filtered to keep positions indicating methylation alone as these are then used to filter out methylation from cfDNA variant calls in a later step. The GATK4 Haplotype Caller was used to call germline variants from germline BAM files with GATK recommended settings. `bcftools mpileup v1.15` was used to generate a pileup file from the cfDNA BAM file. To identify potential variants in the cfDNA sample an extensive filtering process was applied to the pile-up. Specifically, the raw cfDNA variant calls were sorted, decomposed and normalized using `bcftools` to ensure consistency and improve downstream analysis accuracy when filtering germline variants from this set. Normalised variant VCF was further sorted and indexed. Germline variants (Haplotype Caller output VCF) and germline methylation calls (filtered TAMER output) were filtered out if present in cfDNA variants. From this, cfDNA methylation calls (filtered TAMER output) were then filtered out. The variants obtained from the previous step underwent quality filtering based on established criteria, such as read depth, variant allele frequency, quality scores, and annotation information, to ensure the reliability and accuracy of the final variant call set. For SNVs, exclusion criteria were as follows: DP > 200 or DP < 50, MQ < 60, MQBZ < -9, RPBZ < -5 or RPBZ > 5, MQ0F > 0, BQBZ < -4, VAF > 0.3 or VAF < 0.03. For INDELs, the following exclusion criteria were used: DP > 200 or DP < 50, VAF > 0.3 or VAF < 0.03. The resulting cfDNA VCF was annotated using `VEP` and converted to MAF format for easier use of annotations. Finally, the non-cancer SCAN samples that did not progress were used for denoising, by generating a merged VCF of all these samples, which was used to remove shared variants from cfDNA VCF for each cancer sample.

### Fragment-based methylation data analysis

Lists of differentially methylated CpGs identified in TCGA were downloaded via SMART for the cancer types in this study. These lists were used to generate BED files of differentially methylated regions by extending the coordinates 100 bp in each direction and merging any of the resulting regions that overlapped. Any regions that contained fewer than 3 differentially methylated CpGs after this process were discarded along with hypomethylated regions, leaving 2113 hypermethylated regions from 5 TCGA studies (COAD, ESCA, KIRC, KIRP, PAAD). We chose to focus on hypermethylation markers, where baseline methylation should be close to zero in healthy individuals and only successful conversion by the TAPS chemistry would result in a positive signal. For each sample any aligned reads that overlapped with the selected methylation marker regions were matched with their mate pair and the entire fragment was considered together. Fragments that overlapped with fewer than three CpGs in any region were discarded and then methylation calling was performed based on TAPS base changes. Fragments were then classified as either originating from a tumour or originating from healthy cells based on the proportion of callable positions that were modified by the TAPS chemistry. For the purposes of this study the threshold for classification as a tumour fragment was set at 80% modification. Finally, the proportion of fragments classified as originating from tumour cells was calculated for each marker. Samples from the SCAN cohort were used to identify markers that were not specific to cancer. Any marker where one or more fragments classified as tumour were found in these sample was excluded, leaving 377 markers for downstream analysis of the cancer cases and healthy controls.

### Statistical analysis for cancer detection

The clean SNV/INDELs, methylation and coverage signals from each cancer plasma sample were compared to the corresponding signals from non-cancer plasma samples. In the case of the coverage data, the signal was first aggregated in each chromosome arm in each sample. The mean and standard deviation of the total coverage signal for each chromosome arm were calculated across all non-cancer controls. These statistics were subsequently used in the calculation of coverage z-scores for each chromosome arm in each cancer sample. For a particular cancer sample, the z-score at chromosome arm k is given by *z*_*k*_ = (*y*_*k*_ − *m*_*k*_)⁄*s*_*k*_, where *y*_*k*_ is the aggregated coverage signal at arm k in the cancer sample, and m_k_ and s_k_ are the mean and standard deviation of the aggregated coverage signal at chromosome k across the non-cancer controls. The sampling distribution of the statistic 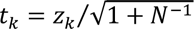 is a t-distribution with *N-1* degrees of freedom, where *N* is the number of non-cancer controls used for the calculation of *m_k_* and *s_k_*. From this, we calculated p-values *p_k_*, which were corrected for multiplicity using the Benjamini-Hochberg procedure. In each cancer sample, those arms with corrected p-values lower than 5% are copy number aberrant in comparison to the corresponding arms of the healthy controls. In order to derive a sample-specific p-value, we selected the aberrant arms, and we combined their p-values into a sample-specific p-value using Stouffer’s method. The cancer samples with p-values less than 5% have copy-number aberrant arms in comparison to the healthy controls. Sample-specific p-values were also derived from the SNV/INDEL and methylation data using the same methodology. In the case of SNVs/INDELs, we use the log_10_ of the mutation burden of each chromosome arm. For the methylation data, we use the logit of the total methylation ratio in each of the 377 methylation markers, derived as explained above. For each sample, a p-value across all three modalities was derived using Stouffer’s method.

### Calculation of ctDNA fractions

In those samples with copy-number aberrations, we were able to calculate the ctDNA fraction based on further analysis of the coverage data. First, the filtered, debiased, denoised and normalised coverage signal was divided into contiguous segments of relatively constant coverage. Genome segmentation was conducted using `DNAcopy v1.68.0` after subsampling the coverage signal every 1000 bins (10^6^ base pairs). Segments less than 3 standard deviations apart were merged into a single segment. Assuming the log_2_-transformed coverage at bin *i* in segment *k* is *y*_*ki*_, we calculated the untransformed coverage *k*_*ki*_ = 2^*yki*+1^. Typically, the plasma cfDNA is a mixture of DNA fragments originating from normal diploid cells and cancer cells. Previous approaches make the strong assumption that the tumour component of the plasma cfDNA originates from one major clone and one subclone. Here, we make the less restrictive assumption that the ctDNA originates from an unspecified number (one or more) of clones of not-necessarily-diploid cancer cells. It follows that the expected value of *k*_*ki*_is equal to 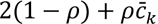 where *ρ* is the ctDNA fraction and 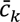 is the average ploidy at segment *k* across all cancer cells. It is important to notice that 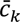 is, in general, not an integer, unless the copy number aberration overlapping segment *k* is a clonal event, i.e., harboured by all cancer cells. We model *k*_*ki*_ using a normal distribution, as follows:

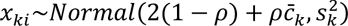

where 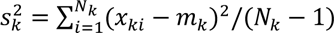 is the observed variance of 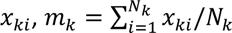 its observed mean, and *N*'_*k*_ the number of bins supporting segment *k*. Since 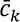 is not an integer, we can impose a uniform prior on it between 0 and a maximum ploidy value, e.g., 4. Similarly, we impose a uniform prior on *ρ* between 0 and 1. The advantage of this formulation is that it allows for a very efficient Gibbs sampling inference scheme for calculating the posterior distribution of 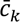 (across all segments) and *ρ*. This scheme consists of repeatedly sampling from the following conditional posteriors for *p* and 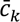:

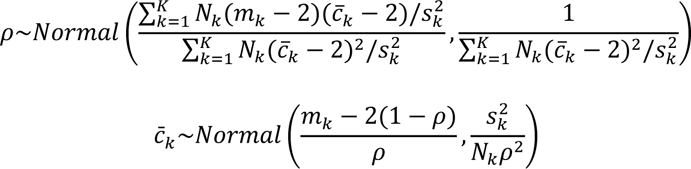

When applied to a particular sample, we ran the above procedure for 10K iterations, and we recorded the mean and variance of the resulting sample chains, after they had attained equilibrium (usually after ∼5K iterations).

### *In silico* data generation for methods validation

In order to validate the above pipeline, we simulated non-cancer and cancer plasma samples at various ctDNA fractions using actual plasma samples as templates. For each *in silico* sample, we simulated aggregate coverage and somatic mutation burden per chromosome arm, and methylation ratios per methylation marker. The aggregate coverage signal in arm *k* for a non-cancer sample was simulated by sampling a point from a normal distribution with mean *m_k_* and variance *s_k_*. These statistics were estimated from the actual non-cancer samples. The aggregate coverage signal in arm *k* for a cancer sample was simulated as a mixture: 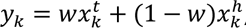, where 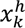 is a random point from a normal distribution with mean *m_k_* and standard deviation 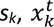 is the aggregate coverage signal in chromosome arm *k* from an actual tumour sample with ctDNA fraction *ρ*, and *w* is the ratio *r/ρ*, where *r* is the target ctDNA fraction of the simulated cancer plasma sample. Mutation burden and methylation signals were generated in the same way using respectively the arm-specific mutation burden and methylation marker-specific statistics in place of *m_k_* and *s_k_* calculated from the healthy controls. For all these mixtures, we used a colorectal cancer plasma sample with *ρ*=9%. For each of ten target ctDNA fractions spanning the range from 0.1% to 2%, we simulated 1000 noncancer and 1000 cancer samples, resulting in a total of 20K simulated plasma samples. The capacity of the previously described statistical methodology to discriminate between cancer and non-cancer plasma samples at decreasing ctDNA fractions was assessed using these simulated data and the area under the ROC curve (AUC) as performance metric.

## Results

For the case cohort with confirmed cancer, the median age was 67.5 years (Table 1). Two-thirds of patients were males. They presented with a wide range of different specific or non-specific symptoms all representative of this patient group. Approximately one-third of patients were classified as Stage 1 or 2 disease with standard TNM staging. Just under two-thirds of patients had colorectal cancer. Follow-up data was available on all patients showing a median overall survival of 8.5 years for both early- and late-stage patients (Fig. S1A). Patients with colorectal cancer had the longest median overall survival (>8.8 years), followed by oesophageal (>8.1 years), ovarian (8 years), renal (3.7 years), breast (>2.3 years) and pancreatic (2 years) cancer patients (Table 1; Fig. S1B).

### Analysis of chromosomal alterations for the detection of ctDNA

Copy number aberrations (large losses or gains of chromosomal material) are considered a hallmark of cancer, manifesting early during tumorigenesis and persisting during subsequent stages of tumour evolution^35–37^. They may involve large regions of each chromosome, chromosomal arms or whole chromosomes (aneuploidies). Such alterations manifest themselves in the data derived from a WGS assay as contiguous upward (gains) or downward (losses) deviations of the number of aligned reads from a baseline corresponding to the non-aberrant (diploid) state. The magnitude of each such deviation depends on the underlying copy number state of the genome at the locus of the aberration, and on the overall tumour fraction in the sequenced sample^36^. To the extent that these aberrations are present in the ctDNA captured and sequenced from the plasma of patients, they can be exploited for the noninvasive detection of almost all cancer types, due to the universal presence of such alternations in the pathogenesis of the disease.

For each cfDNA sample, we divided the genome in 1kb-long non-overlapping bins and counted the number of proper, high-quality alignments that overlapped each bin, followed by a thorough filtering process for removing potential artefacts (see Methods). Since next-generation sequencing and variable mapping can introduce characteristic biases, which alter the chromosomal representation of the original genomic DNA depending on local GC content and mappability^38^, we further applied a statistical approach to simultaneously remove these two types of bias from the number of reads in each bin (see Methods; Fig. 2Ai).

**Figure 2:**
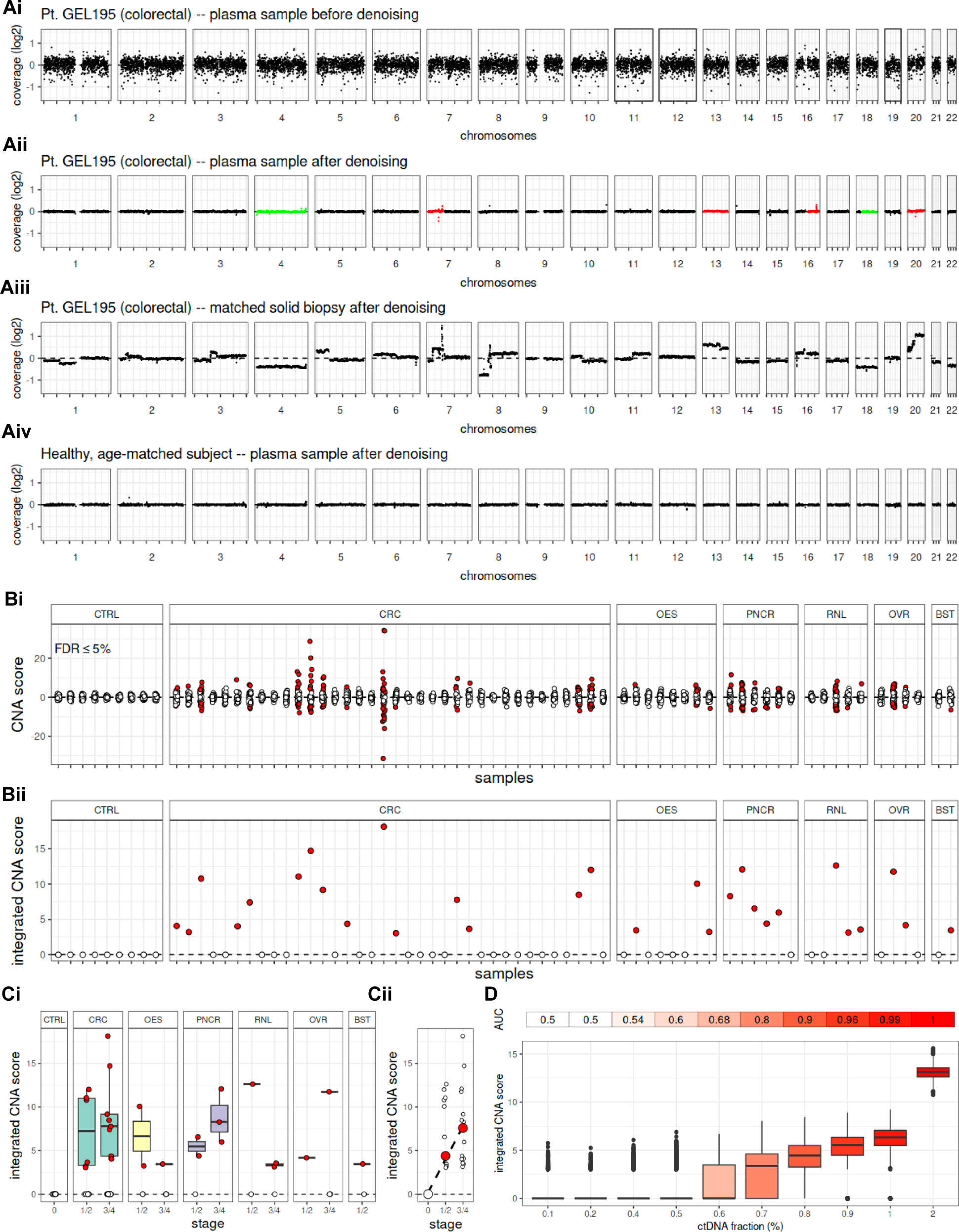
Analysis of copy number aberrations (CNA). A) Coverage signal for patient GEL195 (colorectal cancer) before denoising (Ai), after denoising (Aii) and in the biopsy after denoising (Aiii). A healthy control is also shown for comparison (Aiv). The aggregated coverage signal in each chromosome arm in the plasma sample is compared against the corresponding arm in a cohort of healthy control plasma samples in search of gains (red) or losses (green). Gains in chromosomes 7, 13, 16, 20 and losses in chromosomes 4 and 18 in the plasma sample of patient GEL195 reflect aberrations in the same chromosomes in the matched biopsy, although this has not been used for guiding the analysis. Bi) Scores quantifying coverage imbalances in the chromosome arms of each cancer plasma sample compared to the healthy plasma controls. In each sample, each circle corresponds to a different chromosome arm. Red circles indicate a gain or loss of chromosomal material. Bii) Integrated CNA scores over all chromosome arms in each plasma sample. Red circles indicate the gain or loss of chromosomal material in the corresponding samples. 29 out of 61 cancer samples were correctly identified (sensitivity 47.5%). C) Integrated CNA score against cancer stage and type (Ci) and correlation between median integrated CNA score and stage (Cii). D) *In silico* assessment of CNA analysis performance at increasing ctDNA fractions. At each ctDNA fraction, we simulated 1000 healthy and 1000 cancer plasma samples using actual healthy and cancer plasma samples as templates (see Methods). The area under the receiver operating characteristic (ROC) curve (AUC) is ≥80% at ctDNA fractions ≥0.7%. **CTRL:** controls; **CRC:** colorectal; **OES:** oesophageal; **PNCR:** pancreatic; **RNL:** renal; **OVR:** ovarian; **BST:** breast

Further systematic bias can be introduced due to experimental (e.g., sample preparation) and biological factors (e.g., replication timing), which may hinder detection of subtle read depth differences across the genome in plasma samples with low cfDNA content. For this reason, we further applied a denoising process to remove any systematic errors (see Methods). First, we characterised the background noise using principal component analysis on a panel of normal plasma samples. These were obtained from 21 patients recruited in the Suspected CANcer (SCAN) pathway, which were confirmed healthy or with a non-cancer diagnosis, and therefore appropriate to characterise any non-cancer-specific unwanted variation. This was followed by removing the background noise from each case sample and, subsequently, applying a Savitzky-Golay smoothing filter, which removes any remaining unwanted variation without distorting the underlying signal (Fig. 2Aii-iv).

In order to imitate an early detection scenario, where solid tumour biopsies are not available, we decided to use matched tumour samples for validation purposes only, and never for guiding the search for copy number aberrations. We focused our analysis on whole chromosome arms in order to confidently distinguish positive signals from any remaining background noise (compare Fig. 2Aii to Fig. 2Aiii). Specifically, the denoised coverage signal in each arm was aggregated in each cancer plasma sample, as well as in each of nine age-matched healthy controls. In the controls, the signal ranged from -0.34 to 0.22, while in the cancer samples, the range of coverage values was twice as wide (-0.71 to 0.42). In order to determine whether the aggregate coverage in each arm in a particular patient deviated significantly from the corresponding measurement in the controls (upward indicating chromosomal gain or downward indicating chromosomal loss), we calculated a z-score for each arm as the number of standard deviations from the mean aggregate coverage of the same arm across the normal controls (see Methods). In the normal controls, absolute z-scores were lower than 2.35, while in the cancer plasma samples absolute z-scores took values as high as 34.2 (Fig. 2Bi). Given these scores, p-values were calculated for each arm using an appropriate test statistic and a t-distribution with 8 degrees of freedom (9 controls minus 1), and they were corrected for multiplicity across all arms in each sample using the Benjamini-Hochberg procedure (see Methods). Corrected p-values less than 0.05 (FDR < 5%) indicated significant loss or gain of chromosomal material in the corresponding arm in comparison to the controls. Chromosomal arm alterations were detected in samples from all six cancer types in our cohort, with the number of altered arms per aberrant sample ranging from 1 to 29 (mean = 8.9, std. dev. = 8.1; Fig. 2Bi). Subsequently, we devised a sample-specific copy number aberration score by integrating the p-values across all aberrant arms in each sample using Stouffer’s method. Overall, we detected aberrant arms in 15/36 colorectal, 3/8 oesophageal, 5/6 pancreatic, 3/5 renal, 2/4 ovarian and 1/2 breast cancer plasma samples resulting in 47.5% sensitivity (Fig. 2Bii). The integrated scores in the aberrant samples ranged from 3.0 to 18.1 (mean = 7.6, std. dev. = 4.1). Since the controls were used as the reference set in the calculation of the z-scores, the integrated CNA scores in these samples were all zero (100% specificity). Aberrant samples covered both early (Stage 1 or 2) and more advanced (Stage 3 or 4) cancer stages (Fig. 2Ci) and the median integrated scores correlated with cancer stage (Fig. 2Cii).

In order to evaluate the capacity of the above approach to discriminate between healthy and cancer plasma samples, we conducted Receiver Operating Characteristic (ROC) analysis on synthetic data generated using actual clinical plasma samples as templates. We examined simulated ctDNA fractions ranging from 0.1% to 2% and for each ctDNA fraction, we simulated 1000 healthy controls and 1000 cancer plasma samples by admixing data from actual healthy plasma samples and an actual colorectal cancer plasma sample with 9% tumour burden (see Methods). For each sample in the synthetic case-control dataset, we calculated sample-specific integrated CNA scores, which were used in the construction of an ROC curve. The area under the ROC curve (AUC) at each simulated ctDNA fraction was used to assess classifier performance. The integrated CNA scores correlated with increasing ctDNA fractions 0.6% or higher, while excellent performance (AUC ≥ 80%) was achieved at ctDNA fractions as low as 0.7% (Fig. 2D).

### Analysis of somatic mutation burden for ctDNA detection

Elucidating the profile of somatic mutations present in the plasma cfDNA has been a major research focus in the clinical study of ctDNA as an emerging biomarker for the detection of cancer and monitoring of disease progression. Towards this aim, the major obstacle has been the need to discriminate tumour-originating SNVs and indels from the much more abundant germline mutations and sequencing errors. In order to overcome this problem, one group of methods focuses on deep targeted sequencing of cancer-type-specific panels and driver genes, combined with error-suppression methodologies^39^. Although potentially extremely sensitive, targeted approaches are constrained by the fact that they only sample a small number of human genome equivalents, possibly leading to an inflated false negative rate. In response, an alternative group of approaches centred around shallow WGS and compendiums of patient-specific somatic mutations to guide the analysis has been proposed, thus replacing depth with breadth of sequencing at the cost of increased sequencing noise^22^.

In order to overcome these limitations, we adopted a deep (at least 80×) WGS approach for sensitive mutation detection without requiring matched biopsy samples to guide the analysis. Each plasma sample was a) paired with a matched germline sample for efficient removal of germline mutations, b) processed with bespoke software, which recognises TAPS changes (mC>T) and differentiates these from C>T variants, c) cleaned up using a thorough filtering pipeline to remove sequencing artefacts, and d) further denoised by removing all variants shared with any of 21 age-matched non-cancer plasma samples recruited in the SCAN pathway. All non-synonymous variants were retained for further downstream analysis.

Each control plasma sample harboured between 125 and 192 somatic mutations (mean = 151.1, std. dev. = 25.3; Fig. 3Ai). As expected, cancer plasma samples harboured on average a higher number of mutations, but also showed higher variability (mean = 425.8, std. dev. = 747.3). Plasma samples from pancreatic cancer patients harboured the highest number of mutations on average (mean = 1771.2, std. dev. = 1770.2), followed by oesophageal (mean = 731.8, std. dev. = 1025.9), breast (mean = 385.5, std. dev. = 268.0), colorectal (mean = 288.0, std. dev. = 417.4) and renal (mean = 252.2, std. dev. = 224.0) cancer samples. Across all cancer plasma samples, 11730 genes harboured at least one somatic mutation. Among these, 8902, 1793 and 341 genes carried 1, 2 or 3 somatic mutations, respectively (Fig. 3Aii). As expected from whole genome data, among all mutated genes, only a small fraction of 493 (4.4%) were reported in COSMIC. Most of these were missense, followed by nonsense and splice site mutations (Fig. 3Aiii and Fig. S2). Most of the other mutations are likely acquired passenger events that we used to identify cancer early.

**Figure 3:**
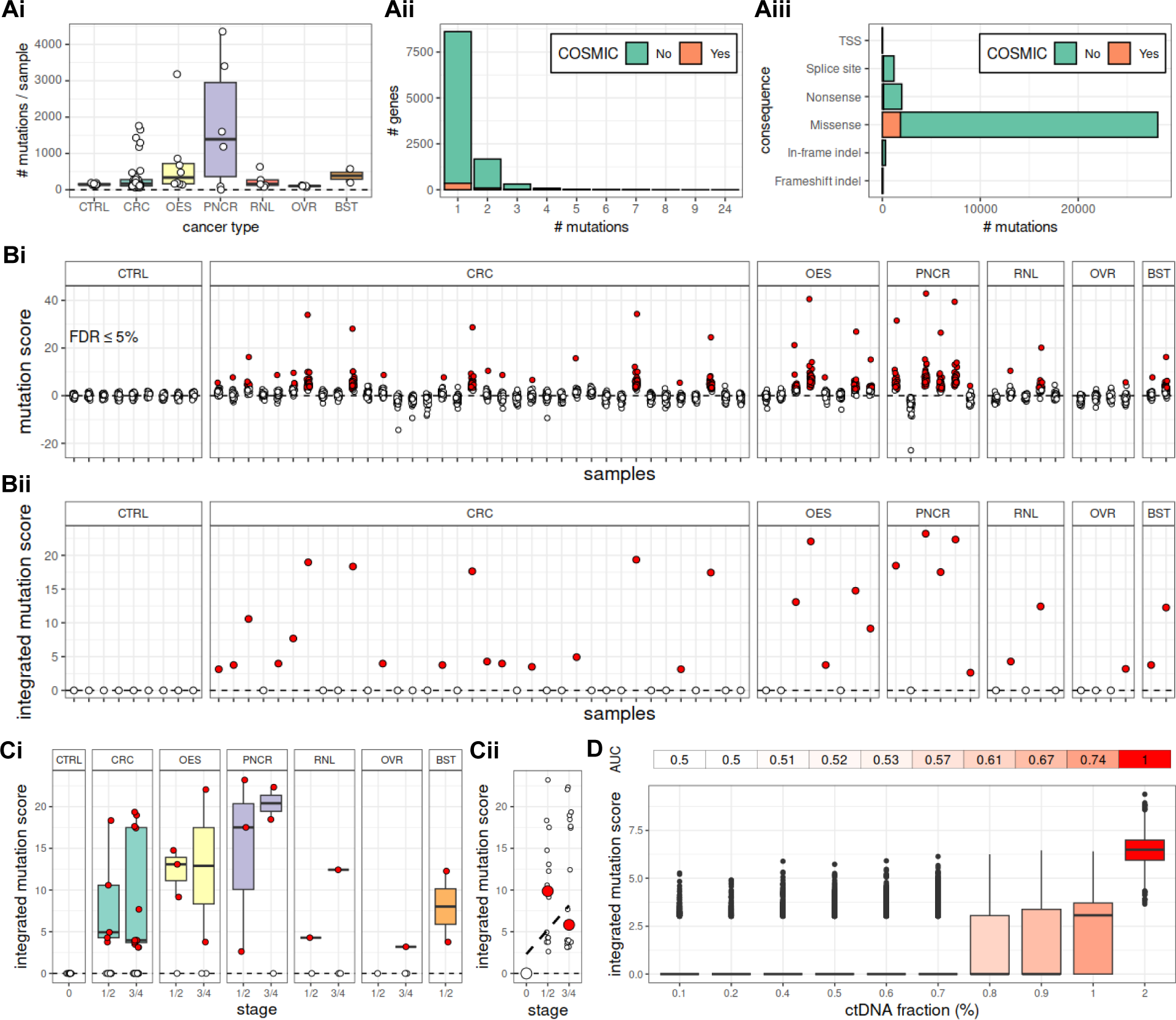
Burden analysis of somatic single nucleotide variants (SNVs) and INDELs (CNA). A) Somatic mutation burden in different cancer types and in healthy controls (Ai), distribution of mutation numbers across genes (Aii) and consequences of mutations (Aiii). In Ai, each circle corresponds to a different plasma sample. Bi) Scores quantifying mutation burden imbalances in the chromosome arms of each cancer plasma sample compared to the healthy plasma controls. In each sample, each circle corresponds to a different chromosome arm. Red circles indicate a difference in the somatic mutations burden of the chromosome arm in relation to the same arm in the controls. Bii) Integrated somatic mutation scores over all chromosome arms in each plasma sample. A red circle indicates a higher mutation burden in the corresponding sample, when compared to the controls. We identified correctly 32 out of 61 cancer plasma samples (sensitivity 52.5%). C) Integrated somatic mutation scores against cancer stage and type (Ci) and correlation between median integrated somatic mutation score and stage (Cii; Spearman’s r = 50%). D) *In silico* validation of somatic mutation analysis at increasing ctDNA fractions. At each ctDNA fraction, we simulated 1000 healthy and 1000 cancer plasma samples using actual healthy and cancer plasma samples as templates (see Methods). The area under the receiver operating characteristic (ROC) curve (AUC) is ≥70% at ctDNA fractions ≥1%. **CTRL:** controls; **CRC:** colorectal; **OES:** oesophageal; **PNCR:** pancreatic; **RNL:** renal; **OVR:** ovarian; **BST:** breast

Next, for each plasma sample, we compared the somatic mutation burden in each chromosome arm against the corresponding arms of nine age-matched healthy controls. Since the density of somatic mutations is not uniform across the whole genome, a comparative analysis based on chromosome arms is expected to be more sensitive, when compared to an analysis based merely on the total somatic mutation burden of the sample. For each chromosome arm in each sample, we calculated the log_10_ of the number of somatic mutations, we derived a zscore as the number of standard deviations from the mean of the same quantity across the controls, and we calculated a p-value, which we corrected for multiplicity across all chromosome arms in each sample using the Benjamini-Hochberg method (see Methods). A corrected p-value lower than 5% indicates a statistically significant difference in the mutation burden of the chromosome arm in comparison to the healthy controls. A significantly increased chromosome arm mutation burden was detected in samples from all cancer types in our cohort, with the number of mutated arms per aberrant sample ranging from 1 to 39 (mean = 13.6, std. dev =15.9; Fig. 3Bi). Subsequently, we devised a sample-specific somatic mutation score by integrating the p-values across all significantly mutated arms in each sample using Stouffer’s method. Overall, we detected arms with increased mutation burden (p-value < 5%) in 17/36 colorectal, 5/8 oesophageal, 5/6 pancreatic, 2/5 renal, 1/4 ovarian and 2/2 breast cancer plasma samples resulting in 52.5% sensitivity (Fig. 3Bii). The integrated scores in the aberrant samples ranged from 2.63 to 23.2 (mean = 10.4, std. dev. = 7.2) and were zero in all healthy controls (100% specificity). Aberrant samples covered both early (Stage 1 or 2) and more advanced (Stage 3 or 4) cancer stages (Fig. 3Ci), while median integrated scores were moderately correlated with cancer stage (Spearman’s r = 50%; Fig. 3Cii). Further evaluation of the discriminatory performance of the above analytical approach using ROC analysis on synthetic data (see Methods) indicates good performance (AUC > 70%) at ctDNA fractions 1% and above (Fig. 3D). This is consistent with a depth of coverage at 100x, which implies a minimum variant allele fraction (VAF) for any mutated locus of 1% (at least one mutated read in a locus covered by 100 reads).

### Analysis of methylation calls for ctDNA detection

DNA methylation is an epigenetic mechanism that can regulate gene expression. When located in a promoter, it typically acts to supress gene expression. Thus, hypomethylation and hypermethylation can lead to increased expression of oncogenes or decreased expression of tumour suppressor genes, respectively. Abnormal DNA methylation patterns are associated with all aspects of cancer pathophysiology, from tumour initiation to tumour progression and metastasis, making DNA methylation abnormalities one of the hallmarks of cancer that can be exploited for disease diagnosis, treatment and monitoring^40–45^.

Current approaches for investigating the methylome using plasma cfDNA^3,43–45^ rely on tissue-specific methylation signatures, which provide a reference set against which the methylation profile of any case of interest is compared to. In this study, we extracted data from several TCGA studies to identify a set of hypermethylated regions, each containing at least three differentially methylated CPGs. We chose to focus on hypermethylation markers, where baseline methylation should be close to zero in healthy individuals and only successful conversion by the TAPS chemistry would result in a positive signal. Subsequently, we identified fragments in each cfDNA sample overlapping at least three CPGs in any of these regions and we calculated the overall methylation level of the fragment. We decided to follow a fragment-rather than a locus-centric approach, since it is known that in low ctDNA fraction settings, this can lead to increased sensitivity^44^. Fragments with higher than 80% methylation were classified as tumour-originating and the total fraction of tumour-originating fragments was calculated in each region. An additional level of denoising was added by removing all regions containing at least one tumour-originating fragment in any of the non-cancer SCAN controls.

Next, we compared the fraction of tumour-originating fragments in each region between each plasma sample and nine age-matched healthy controls. For each region in each sample, we calculated the logit of the fragment fraction, we derived a z-score and a corresponding p-value, which was corrected for multiplicity across all regions in each sample using the Benjamini-Hochberg method (see Methods). The number of regions with significantly higher burden of tumour-originating fragments per sample ranged from 1 to 371 (mean = 113.6, std. dev. 134.1; Fig. 4Ai) and, from the p-values of these regions, an integrated methylation score was derived for each sample using Stouffer’s method. Overall, we detected regions with significantly higher methylation burden (p-value < 5%) in 18/36 colorectal, 2/8 oesophageal, 1/6 pancreatic, 4/5 renal, 3/4 ovarian and 0/2 breast cancer plasma samples resulting in 45.9% sensitivity (Fig. 4Aii). The integrated methylation scores in the aberrant samples ranged from 3.8 to 64.0 (mean = 24.5, std. dev. = 17.7) and were zero in all healthy controls (100% specificity). Aberrant samples covered both early (Stage 1 or 2) and more advanced (Stage 3 or 4) cancer stages (Fig. 4Bi) and the median integrated scores correlated with cancer stage (Fig. 4Bii). ROC analysis on synthetic data (see Methods) indicated good discriminatory capacity of the above methodology (AUC > 70%) at ctDNA fractions 0.8% and above (Fig. 3C).

**Figure 4:**
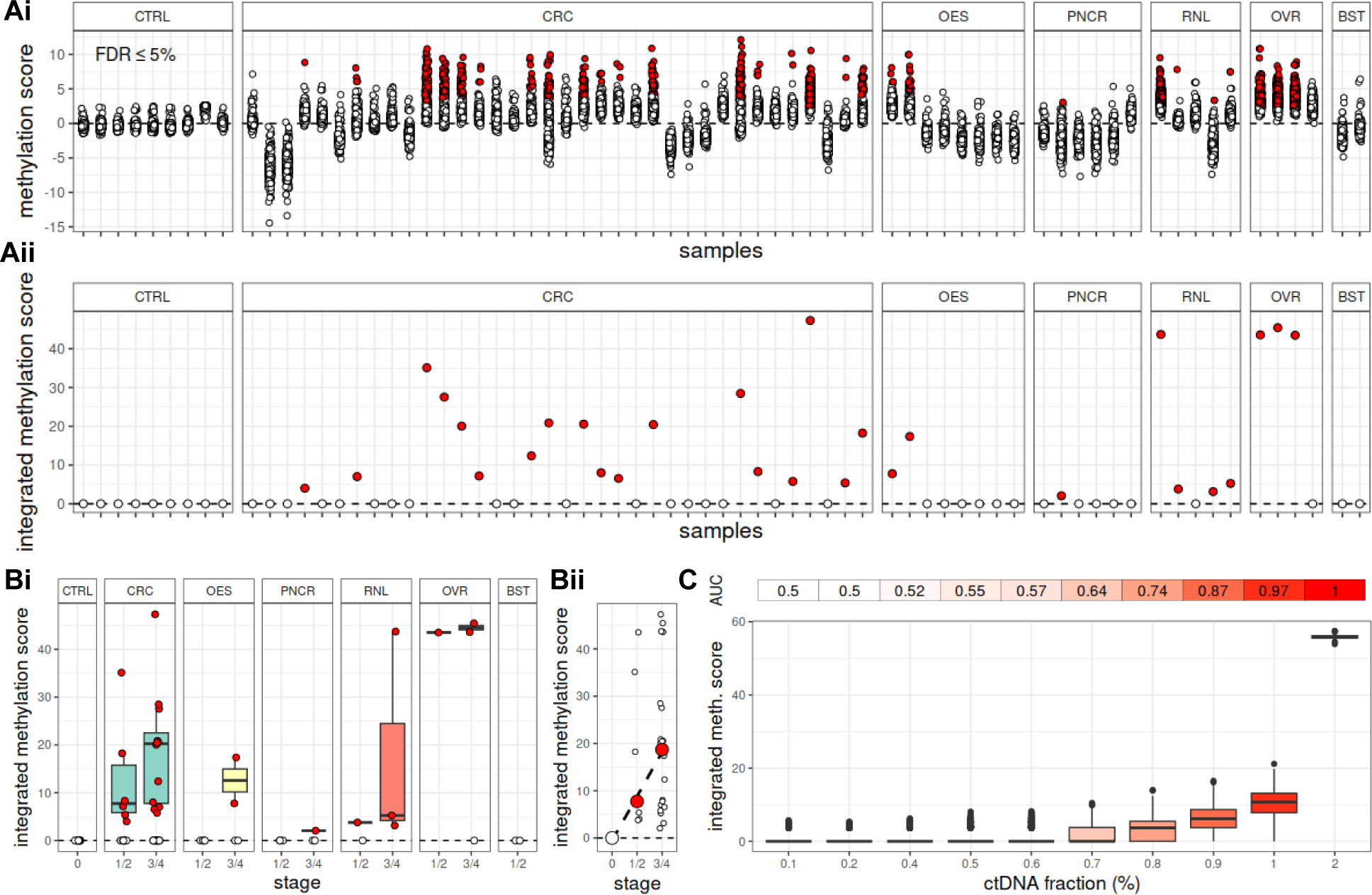
Overview of methylation analysis. Ai) Scores quantifying imbalances in the methylation burden in any of 377 regions (extracted from TCGA; see Methods) in each cancer plasma sample compared to the healthy plasma controls. Each circle corresponds to a different region and red circles indicate over-methylation of the corresponding regions between the cancer plasma and the controls. Aii) Integrated methylation scores over all regions in each plasma sample. A red circle indicates an over-methylated plasma sample, when compared to the controls. We identified correctly 28 out of 61 cancer plasma samples, which corresponds to a 45.9% sensitivity. B) Integrated methylation scores against cancer stage and type (Bi) and correlation between median integrated methylation scores and stage (Bii). C) *In silico* validation of methylation analysis at increasing ctDNA fractions. At each ctDNA fraction, we simulated 1000 healthy and 1000 cancer plasma samples using actual healthy and cancer plasma samples as templates (see Methods). The area under the receiver operating characteristic (ROC) curve (AUC) is ≥70% at ctDNA fractions ≥0.8%. **CTRL:** controls; **CRC:** colorectal; **OES:** oesophageal; **PNCR:** pancreatic; **RNL:** renal; **OVR:** ovarian; **BST:** breast

### Integration of multiple genomic modalities for ctDNA detection

The genomic data modalities analysed above provide three independent and complementary assessments of tumour content in the plasma. We reasoned that combining these data modalities may enrich the available signal and increase the sensitivity of detection of ctDNA in each plasma sample, particularly in cases where not all three types of abnormalities are detectable due to the sparsity of ctDNA. In each sample, given the sample-specific p-values for each data modality, an integrated multimodal score and corresponding p-value can be readily calculated using Stouffer’s method. Although in this study all three modalities are combined using equal weights, unequal weights specific to each genomic data type can also be introduced, if necessary. Furthermore, this approach is applicable even if not all three modalities are available in a particular sample. In our cohort, multimodal scores across aberrant plasma samples ranged from 1.8 to 36.5 (mean = 11.8, std. dev. = 9.8) and they were all zero across the controls (specificity 100%). Significant p-values (<5%) indicating the presence of ctDNA were calculated in 29/36 colorectal, 7/8 oesophageal, 5/6 pancreatic, 5/5 renal, 4/4 ovarian and 2/2 breast cancer plasma samples resulting in a substantially increased sensitivity of 85.2%, compared to the sensitivity of each independent data modality (Fig. 5A). Positive results covered both early (stages 1 or 2) and late (stages 3 and 4) cancer cases (Fig. 5Bi), and median multimodal scores correlated with cancer stage (Fig. 5Bii), while ROC analysis using synthetic data indicated excellent performance (AUC ≥ 80%) at ctDNA fractions as low as 0.7% (Fig. 5C).

**Figure 5:**
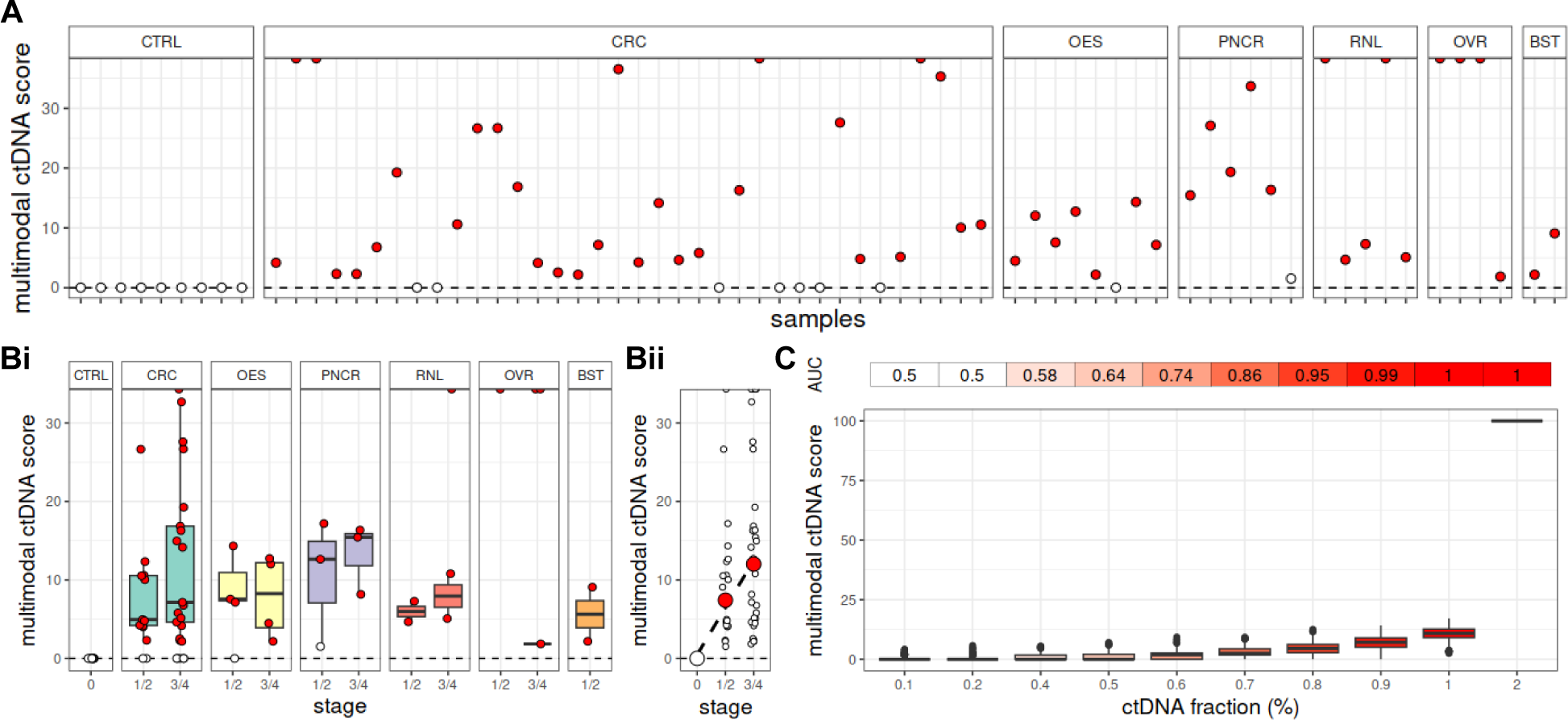
Integration of genomic data modalities for ctDNA detection. A) Multimodal scores for the quantification of plasma ctDNA generated from the integration of copy number aberrations, somatic SNVs and INDELs, and methylation signals in each plasma sample. A red circle indicates a higher ctDNA burden in comparison to the healthy controls. 52 out of 61 cancer plasma samples were correctly identified as such, which corresponds to 85.2% sensitivity. This is higher than the sensitivity of any of the three data modalities. B) Multimodal scores against cancer stage and type (Bi) and correlation between median multimodal scores and stage (Bii). C) *In silico* validation of multimodal analysis at increasing ctDNA fractions. At each ctDNA fraction, we simulated 1000 healthy and 1000 cancer plasma samples using actual healthy and cancer plasma samples as templates (see Methods). The area under the receiver operating characteristic (ROC) curve (AUC) is ≥80% at ctDNA fractions ≥0.7%. **CTRL:** controls; **CRC:** colorectal; **OES:** oesophageal; **PNCR:** pancreatic; **RNL:** renal; **OVR:** ovarian; **BST:** breast

### Multimodal ctDNA detection for post-operative MRD and adjuvant therapy response tracking in colorectal cancer without matched tumour

Up to this point, we focused our analysis on treatment-naive and verified non-cancer plasma samples, aiming to imitate an early detection scenario. In order to evaluate our approach for tracking disease progression^46–49^, we further analysed preand post-operative plasma samples from 10 colorectal cancer patients with detectable ctDNA in the pre-operative sample, half of which had also received adjuvant therapy following surgical resection. For each plasma sample in each patient, integrated ctDNA scores and p-values from all three data modalities were calculated as for the pre-operative samples. A p-value threshold of 5% was taken to indicate the presence of ctDNA in the plasma. As before, matched tumour biopsies, if available, were not used for guiding the analysis, since this carries the risk of missing recurrence due to clonal evolution of the primary tumour or the presence of second primary. Besides, it has been shown that in the real-world, there are significant delays in accessing tumour tissue biopsies for monitoring.

Five patients did not receive adjuvant therapy. In one of them, (GEL193; Fig. 6A), ctDNA was detectable in the plasma 1 year after surgery. The patient was diagnosed with inoperable metastatic rectal cancer three years later and a possible lung adenocarcinoma on radiological examination. Two patients (GEL066, Fig. S3Ai; GEL339, Fig. S3Aii) had detectable ctDNA in the plasma 2 years and 6 months after surgery, respectively. They were found to have tubular adenomas with low-grade dysplasia on routine follow-up 2 years and 7 months after the last post-surgery blood samples were collected, respectively. The remaining two patients permanently cleared ctDNA 3 months and 16 months after surgery, respectively (Fig. S3Aiii,iv). One of them (GEL107) is alive 8 years after the last blood sample was taken, while the other (GEL197) died from sudden cardiac death after 5 years.

**Figure 6:**
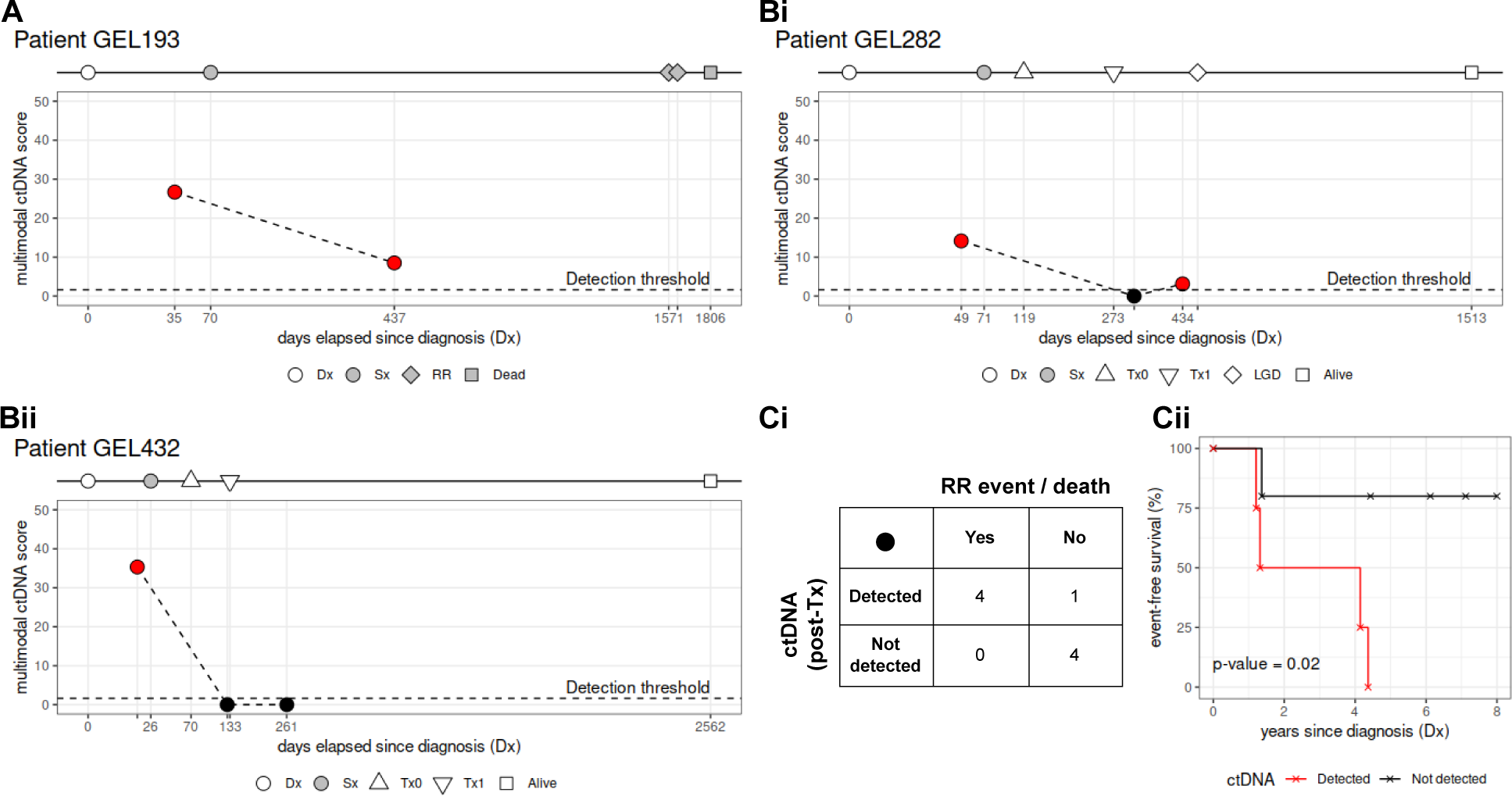
Multimodal ctDNA detection for post-operative MRD and adjuvant therapy response tracking in colorectal cancer without matched tumour. A) Tracking post-operative MRD in case GEL193. ctDNA was detectable in the plasma 1 year after surgery and this correlated with inoperable metastatic rectal cancer and a possible lung adenocarcinoma suggested through radiological examination, both of which were recorded approximately 3 years after the post-operative plasma sample was collected. B) Tracking response to adjuvant therapy following surgery. Case GEL282 (Bi) did not have detectable ctDNA immediately after the last cycle of treatment. However, low ctDNA burden was detected 5 months later, which correlated with the presence of tubular adenomas with low grade dysplasia in the sigmoid colon at around the same time. Case GEL432 (Bii) did not have detectable ctDNA shortly after the last cycle of treatment and was still alive approximately 6 years after the last plasma sample was collected. C) Confusion matrix (Ci) and event-free (i.e., no recurrence, metastasis, or precancerous adenomas present) survival (Cii) in 9 patients with colorectal cancer. In 8 out 9 patients, ctDNA burden after the end of surgery and/or adjuvant therapy correlated with the presence/absence of clinical events, such as recurrence or pre-cancerous adenomas (Ci). Absence of ctDNA detection after the end of surgery/adjuvant treatment correlated with longer survival times (Cii). **Dx:** diagnosis; **Sx:** surgery; **Tx0:** first cycle of adjuvant therapy; **Tx1:** last cycle of adjuvant therapy; **RR:** recurrence; **LGD:** low-grade dysplasia

Five patients received adjuvant therapy after surgery. GEL282 did not have detectable ctDNA immediately after the end of adjuvant treatment (Fig. 6Bi). However, a low positive ctDNA burden was detected 5 months later, which correlated with the presence of tubular adenomas with low grade dysplasia in the sigmoid colon at around the same time. Cases GEL432 (Fig. 6Bii) and GEL205 (Fig. S3Bi) did not have detectable ctDNA after the end of treatment and they were both alive 6 years and 4 years after the last plasma sample was collected, respectively.

For case GEL195 (Fig. S3Bii), a post-operative blood sample collected 3 months after surgery indicated the presence of ctDNA in the plasma, although there was a 74% reduction in tumour burden compared to the pre-surgery sample (multimodal ctDNA score before surgery: 16.86, after surgery: 4.34). Since the sample was taken 1½ months after the first cycle and approximately 4 months before the last cycle of adjuvant treatment, we presume that there would be no detectable ctDNA in the plasma after the end of therapy. The patient was still alive approximately 8 years after the last blood sample was collected. Finally, case GEL223 (Fig. S3Biii) did not show detectable ctDNA levels 8 months after the last cycle of adjuvant therapy, despite the diagnosis of prostate adenocarcinoma 1 month earlier. This was treated with hormone therapy and radiotherapy, and the patient was still alive 6½ years after the last plasma sample was collected.

Overall, presence or absence of ctDNA in the last plasma sample was correlated with adjuvant therapy or clinical outcome in 9 out of 10 cases (the exception was case GEL223). Furthermore, in the cases for which we had plasma samples after surgery or after the end of adjuvant therapy (n = 9 cases), event-free survival (i.e., no recurrence, metastasis or pre-cancerous adenomas) was correlated with the absence of detectable ctDNA posttreatment (two-sided log-rank test; p-value = 0.02; Fig. 6Ci, ii).

## Discussion

To our knowledge, this is first study to explore whether comprehensive multimodality (CNV, SNV and methylation modifications) WGS of ctDNA at 80× or higher can improve the detection of cancer signals from liquid biopsies. Current choices of methodologies are mainly driven by cost and have focussed on targeted deep sequencing of a small number of targets or single modalities. By contrast, after sequencing ctDNA at a coverage depth of at least 80×, we combined genome-wide information from acquired CNAs, SNVs and methylation changes with a highly effective de-noising strategy to reveal cancer signals to high sensitivity and specificity.

Current methods for methylation detection from liquid biopsies employ bisulfite treatment^3,50,51^ or methylation specific PCR^52^. Detection of methylated cytosines from TET-Assisted Pyridine Borane Sequencing (TAPS) has been shown to compare favourably against bisulfite treatment. In contrast to harsh chemicals like bisulfite, TAPS does not destroy low-abundance ctDNA. Moreover, it only converts the 5% of methylated cytosines as opposed to the 95% of unmethylated cytosines, thereby preserving the genetic code. Here, to increase the sensitivity of cancer signal detection from CNAs, we employ this technology to reveal acquired SNVs from the same sample and sequencing run using an in-house bioinformatics tool called TAMER to correct for C>T changes. As we only had paired tissue samples available for a minority of cases, future studies are needed to understand whether the mutation spectrum seen in plasma using this approach is representative of that seen in corresponding tissue biopsies.

This is a diagnostic accuracy study using a case-control design and samples from patients presenting with symptoms of cancer who were subsequently referred to surgery with curative intent. In order to define the true positive and negative predictive value (PPV; NPV) in this cohort of patients, a much larger study on thousands of patients would be needed, similar to the SYMPLIFY study that recruited over 6000 patients presenting with non-specific symptoms of cancer to rapid diagnostic pathways in the UK’s National Health Service^6^. If we apply a cancer prevalence of 6.7% observed in SYMPLIFY to model our test performance and assume a pre-set test sensiwvity of 85.2% (which is the overall sensiwvity we observed in our study, when the specificity is fixed at 100%, based on 9 negative controls), then the NPV would be 98.9% in this group of pawents and arguably sufficient to rule out cancer in a primary care seyng where the likelihood of cancer is low.

Further refinement of the technology by integrating fragment size distribution, mutational signatures, telomere length and other genome-wide cancer signals is ongoing. It remains to be seen whether these increase sensitivity and specificity further. As we have generated whole (epi-)genome data, tissue of origin (TOO) prediction could also be evaluated without having to repeat experiments, but it remains to be seen whether TOO prediction is useful in clinical practice or whether it leads to an increase in false positive cancer calls. We therefore decided to exclude it from the current analysis.

In conclusion, we show that our approach to multi-modality analysis of ctDNA using deep whole genome sequencing combined with TAPS detects cancer signals in early-stage cancer to high accuracy. The next step will be to perform a prospective study in unselected consecutive high-risk individuals to establish its predictive values.

## Data Availability

All data produced in the present study are available upon reasonable request to the authors

## Acknowledgements

This research was funded by Innovate UK and the National Institute for Health Research (NIHR) Oxford Biomedical Research Centre. We further acknowledge the contribution to this study made by the Oxford Centre for Histopathology Research and the Oxford Radcliffe Biobank, which are supported by the University of Oxford, the Oxford CRUK Cancer Centre and the NIHR Oxford Biomedical Research Centre (Molecular Diagnostics Theme/Multimodal Pathology Subtheme), and the NIHR CRN Thames Valley network. The views expressed are those of the authors and not necessarily those of the NHS, the NIHR or the Department of Health and Social Care.

---

**Figure S1:**
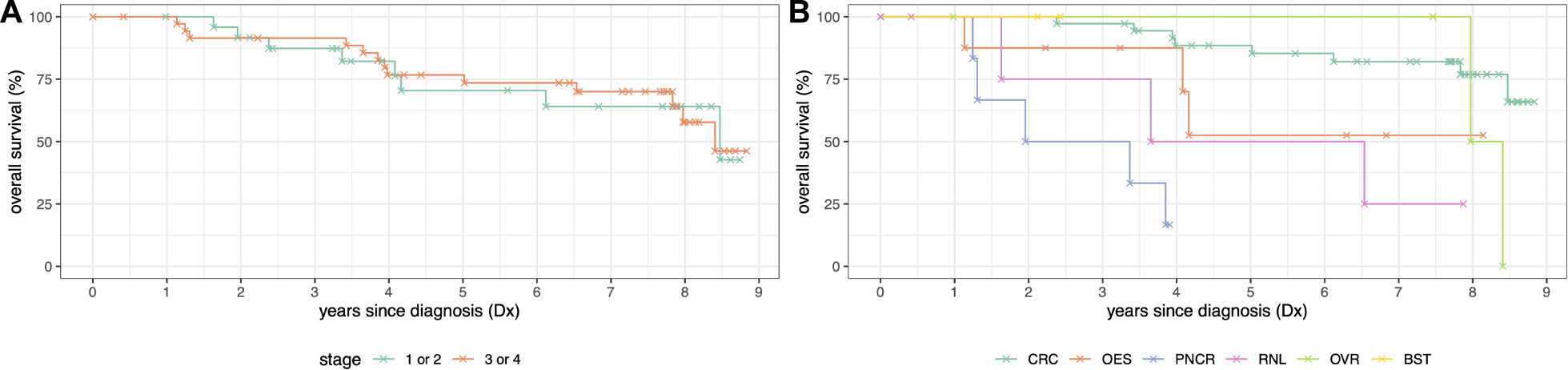
Overall survival by stage and cancer site. A) Overall survival of cases with confirmed cancer by stage. Median overall survival for both early-stage (1 or 2) and late-stage (3 or 4) disease is approximately 8.5 years since diagnosis. B) Overall survival by cancer type. Patients with colorectal cancer had the longest median overall survival (>8.8 years), followed by oesophageal (>8.1 years), ovarian (8 years), renal (3.7 years), breast (>2.3 years) and pancreatic (2 years) cancer patients. **CTRL:** controls; **CRC:** colorectal; **OES:** oesophageal; **PNCR:** pancreatic; **RNL:** renal; **OVR:** ovarian; **BST:** breast

**Figure S2:**
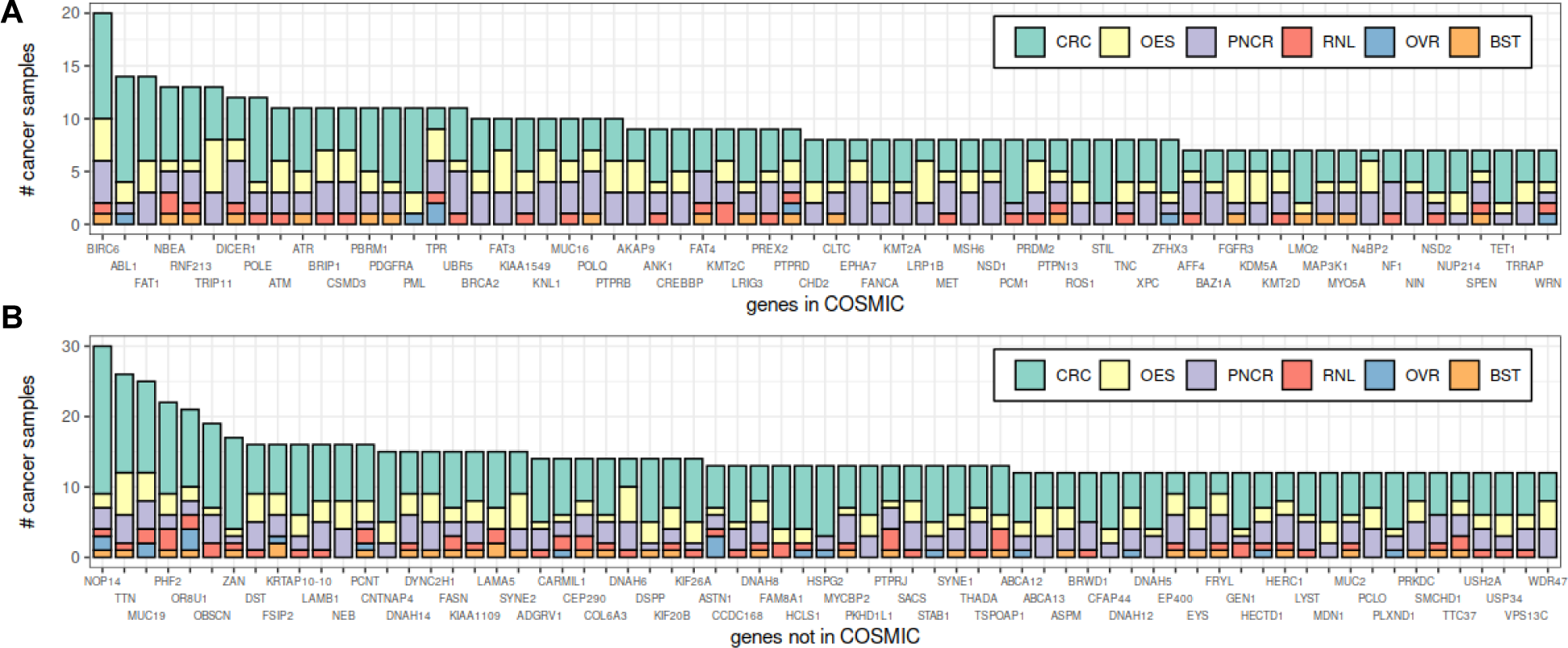
Frequency of mutated genes. Number of samples mutated in genes found (A) and not found (B) in COSMIC. Only the top 66 genes are shown in each panel, due to space restrictions. **CTRL:** controls; **CRC:** colorectal; **OES:** oesophageal; **PNCR:** pancreatic; **RNL:** renal; **OVR:** ovarian; **BST:** breast

**Figure S3:**
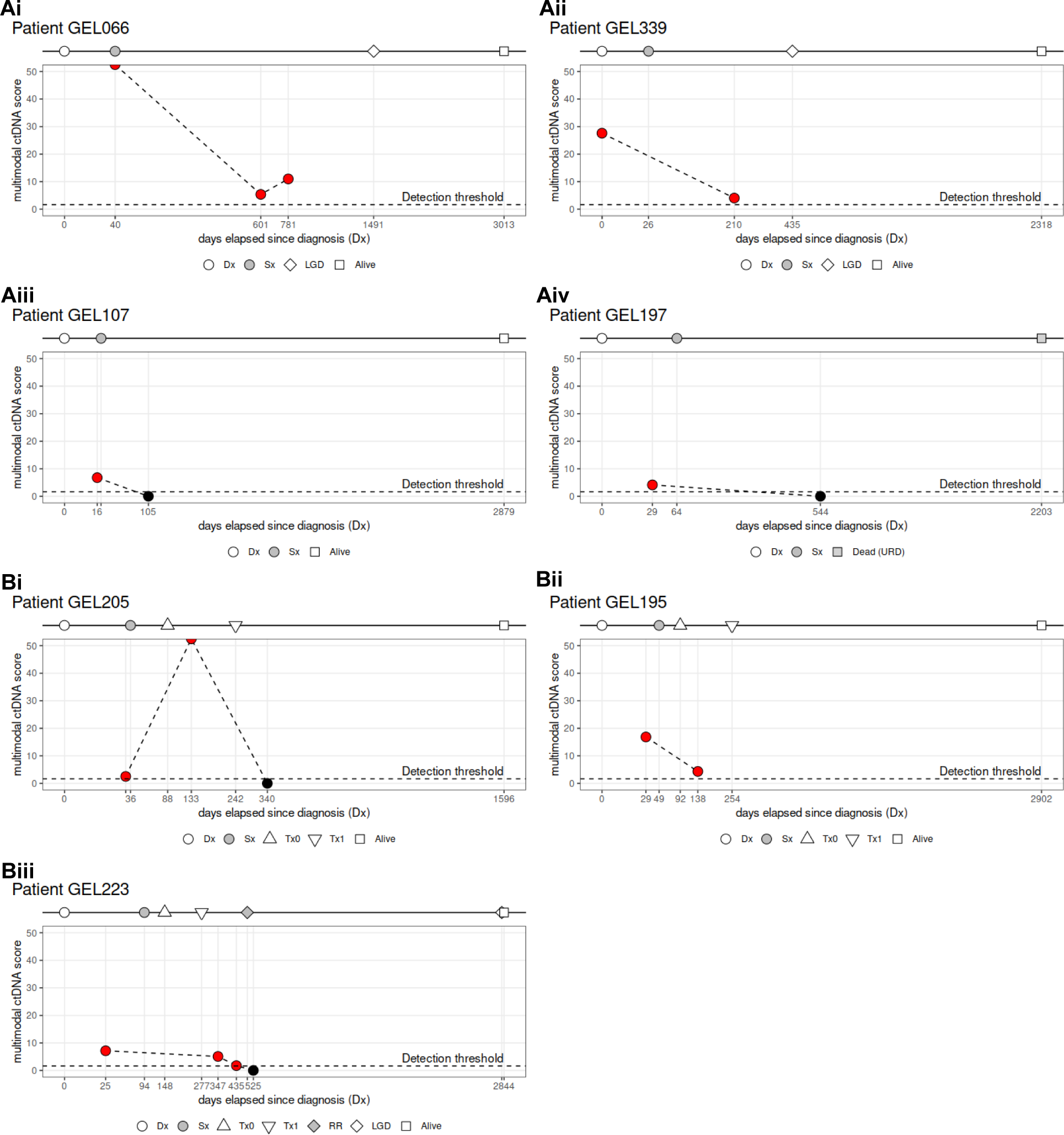
Multimodal ctDNA detection for post-operative MRD and adjuvant therapy response tracking in colorectal cancer without matched tumour. A) Surgical patients who did not receive adjuvant treatment. Detection of ctDNA at the last post-surgery plasma sample correlated with the presence of pre-cancerous adenomas (Ai, Aii). Absence of detection of ctDNA at the last post-surgery plasma sample correlated with progression-free survival (Aiii, Aiv). Notice that case GEL197 died suddenly from unrelated reasons, 5 years after the last blood sample was collected. B) Surgical patients who received adjuvant treatment. In case GEL205 (Bi), a relatively high transient ctDNA burden was detected during adjuvant treatment, but there was no detectable ctDNA shortly after the last cycle of treatment and the patient was alive 4 years later. For case GEL195 (Bii), a post-operative blood sample collected 3 months after surgery indicated the presence of ctDNA in the plasma, although there was a 74% reduction in tumour burden compared to the pre-surgery sample (multimodal ctDNA score before surgery: 16.86, after surgery: 4.34). Since the sample was taken 1½ months after the first cycle and approximately 4 months before the last cycle of adjuvant treatment, we presume that there would be no detectable ctDNA in the plasma after the end of therapy. Case GEL223 (Biii) did not show detectable ctDNA levels 8 months after the last cycle of adjuvant therapy, despite the diagnosis of prostate adenocarcinoma 1 month earlier. This was treated with hormone therapy and radiotherapy, and the patient was still alive 6½ years after the last plasma sample was collected. **Dx:** diagnosis; **Sx:** surgery; **Tx0:** first cycle of adjuvant therapy; **Tx1:** last cycle of adjuvant therapy; **RR:** recurrence; **LGD:** low-grade dysplasia; **URD:** unrelated death

## References

1. Crosby, D. et al. Early detection of cancer. Science 375, eaay9040 (2022).

2. Lennon, A. M. et al. Feasibility of blood testing combined with PET-CT to screen for cancer and guide intervention. Science 369, (2020).

3. Liu, M. C. et al. Sensitive and specific multi-cancer detection and localization using methylation signatures in cell-free DNA. Ann. Oncol. 31, 745–759 (2020).

4. Neal, R. D. et al. Cell-Free DNA-Based Multi-Cancer Early Detection Test in an Asymptomatic Screening Population (NHS-Galleri): Design of a Pragmatic, Prospective Randomised Controlled Trial. Cancers 14, (2022).

5. Nadauld, L. D. et al. The PATHFINDER Study: Assessment of the Implementation of an Investigational Multi-Cancer Early Detection Test into Clinical Practice. Cancers 13, (2021).

6. Nicholson, B. D. et al. Multi-cancer early detection test in symptomatic patients referred for cancer investigation in England and Wales (SYMPLIFY): a large-scale, observational cohort study. Lancet Oncol. 24, 733–743 (2023).

7. Lyratzopoulos, G., Wardle, J. & Rubin, G. Rethinking diagnostic delay in cancer: how difficult is the diagnosis? BMJ 349, g7400 (2014).

8. Moore, S. F., Price, S. J., Chowienczyk, S., Bostock, J. & Hamilton, W. The impact of changing risk thresholds on the number of people in England eligible for urgent investigation for possible cancer: an observational cross-sectional study. Br. J. Cancer 125, 1593–1597 (2021).

9. CancerData. https://www.cancerdata.nhs.uk/.

10. Chapman, D. et al. First results from five multidisciplinary diagnostic centre (MDC) projects for non-specific but concerning symptoms, possibly indicative of cancer. Br. J. Cancer 123, 722–729 (2020).

11. Chapman, D. et al. Non-specific symptoms-based pathways for diagnosing less common cancers in primary care: a service evaluation. Br. J. Gen. Pract. 71, e846–e853 (2021).

12. Fitzgerald, R. C., Antoniou, A. C., Fruk, L. & Rosenfeld, N. The future of early cancer detection. Nat. Med. 28, 666–677 (2022).

13. Hamilton, W., Walter, F. M., Rubin, G. & Neal, R. D. Improving early diagnosis of symptomatic cancer. Nat. Rev. Clin. Oncol. 13, 740–749 (2016).

14. Rubin, G. et al. The expanding role of primary care in cancer control. Lancet Oncol. 16, 1231–1272 (2015).

15. Nicholson, B. D. & Lyratzopoulos, G. Progress and priorities in reducing the time to cancer diagnosis. British journal of cancer vol. 128 468–470 (2023).

16. Brito-Rocha, T., Constâncio, V., Henrique, R. & Jerónimo, C. Shifting the Cancer Screening Paradigm: The Rising Potential of Blood-Based Multi-Cancer Early Detection Tests. Cells 12, (2023).

17. Cohen, J. D. et al. Detection and localization of surgically resectable cancers with a multi-analyte blood test. Science 359, 926–930 (2018).

18. Phallen, J. et al. Direct detection of early-stage cancers using circulating tumor DNA. Sci. Transl. Med. 9, (2017).

19. Killcoyne, S. et al. Genomic copy number predicts esophageal cancer years before transformation. Nat. Med. 26, 1726–1732 (2020).

20. Cristiano, S. et al. Genome-wide cell-free DNA fragmentation in patients with cancer. Nature 570, 385–389 (2019).

21. Bao, H., et al. Letter to the Editor: An ultra-sensitive assay using cell-free DNA fragmentomics for multicancer early detection. Mol. Cancer 21, 129 (2022).

22. Zviran, A. et al. Genome-wide cell-free DNA mutational integration enables ultra-sensitive cancer monitoring. Nat. Med. (2020) doi:10.1038/s41591-020-0915-3.

23. Widman, A. J. et al. Machine learning guided signal enrichment for ultrasensitive plasma tumor burden monitoring. bioRxiv 2022.01.17.476508 (2022) doi:10.1101/2022.01.17.476508.

24. Peneder, P. et al. Multimodal analysis of cell-free DNA whole-genome sequencing for pediatric cancers with low mutational burden. Nat. Commun. 12, 3230 (2021).

25. Cutts, A. et al. Characterisation of the changing genomic landscape of metastatic melanoma using cell free DNA. NPJ Genom Med 2, 25 (2017).

26. Li, Y. et al. Multi-omics integrated circulating cell-free DNA genomic signatures enhanced the diagnostic performance of early-stage lung cancer and postoperative minimal residual disease. EBioMedicine 91, 104553 (2023).

27. Ganesamoorthy, D. et al. Whole genome deep sequencing analysis of cell-free DNA in samples with low tumour content. BMC Cancer 22, 85 (2022).

28. Grunau, C., Clark, S. J. & Rosenthal, A. Bisulfite genomic sequencing: systematic investigation of critical experimental parameters. Nucleic Acids Res. 29, E65–5 (2001).

29. Liu, Y. et al. Bisulfite-free direct detection of 5-methylcytosine and 5-hydroxymethylcytosine at base resolution. Nat. Biotechnol. 37, 424–429 (2019).

30. Liu, Y. et al. Accurate targeted long-read DNA methylation and hydroxymethylation sequencing with TAPS. Genome Biol. 21, 54 (2020).

31. Vaisvila, R. et al. Enzymatic methyl sequencing detects DNA methylation at single-base resolution from picograms of DNA. Genome Res. 31, 1280–1289 (2021).

32. Siejka-Zielińska, P. et al. Cell-free DNA TAPS provides multimodal information for early cancer detection. Sci Adv 7, eabh0534 (2021).

33. Hamilton, W., Hajioff, S., Graham, J. & Schmidt-Hansen, M. Suspected cancer (part 2--adults): reference tables from updated NICE guidance. BMJ 350, h3044 (2015).

34. Savitzky, A. & Golay, M. J. E. Smoothing and Differentiation of Data by Simplified Least Squares Procedures. Anal. Chem. 36, 1627–1639 (1964).

35. Leary, R. J. et al. Detection of chromosomal alterations in the circulation of cancer patients with wholegenome sequencing. Sci. Transl. Med. 4, 162ra154 (2012).

36. Adalsteinsson, V. A. et al. Scalable whole-exome sequencing of cell-free DNA reveals high concordance with metastatic tumors. Nat. Commun. 8, 1324 (2017).

37. Steele, C. D. et al. Signatures of copy number alterations in human cancer. Nature 606, 984–991 (2022).

38. Benjamini, Y. & Speed, T. P. Summarizing and correcting the GC content bias in high-throughput sequencing. Nucleic Acids Res. 40, e72 (2012).

39. Newman, A. M. et al. Integrated digital error suppression for improved detection of circulating tumor DNA. Nat. Biotechnol. 34, 547–555 (2016).

40. Esteller, M. Epigenetics in cancer. N. Engl. J. Med. 358, 1148–1159 (2008).

41. Sharma, S., Kelly, T. K. & Jones, P. A. Epigenetics in cancer. Carcinogenesis 31, 27–36 (2010).

42. Markowitz, S. D. & Bertagnolli, M. M. Molecular origins of cancer: Molecular basis of colorectal cancer. N. Engl. J. Med. 361, 2449–2460 (2009).

43. Kang, S. et al. CancerLocator: non-invasive cancer diagnosis and tissue-of-origin prediction using methylation profiles of cell-free DNA. Genome Biol. 18, 53 (2017).

44. Li, W. et al. CancerDetector: ultrasensitive and non-invasive cancer detection at the resolution of individual reads using cell-free DNA methylation sequencing data. Nucleic Acids Res. 46, e89 (2018).

45. Choy, L. Y. L. et al. Single-Molecule Sequencing Enables Long Cell-Free DNA Detection and Direct Methylation Analysis for Cancer Patients. Clin. Chem. 68, 1151–1163 (2022).

46. Tie, J. et al. Circulating Tumor DNA Analysis Guiding Adjuvant Therapy in Stage II Colon Cancer. N. Engl. J. Med. 386, 2261–2272 (2022).

47. Kotani, D. et al. Molecular residual disease and efficacy of adjuvant chemotherapy in patients with colorectal cancer. Nat. Med. 29, 127–134 (2023).

48. Abbosh, C. et al. Tracking early lung cancer metastatic dissemination in TRACERx using ctDNA. Nature 616, 553–562 (2023).

49. Killock, D. Early MRD predicts disease recurrence and benefit from adjuvant chemotherapy in CRC. Nature reviews. Clinical oncology vol. 20 137 (2023).

50. Klein, E. A. et al. Clinical validation of a targeted methylation-based multi-cancer early detection test using an independent validation set. Ann. Oncol. 32, 1167–1177 (2021).

51. Jamshidi, A. et al. Evaluation of cell-free DNA approaches for multi-cancer early detection. Cancer Cell 40, 1537–1549.e12 (2022).

52. Parikh, A. R. et al. Minimal Residual Disease Detection using a Plasma-only Circulating Tumor DNA Assay in Patients with Colorectal Cancer. Clin. Cancer Res. 27, 5586–5594 (2021).

